# Neuromuscular risk factors for non-contact knee injury: a systematic review and meta-analysis

**DOI:** 10.1101/2021.09.06.21263164

**Authors:** Bonar McGuire, Ben King

## Abstract

**Objectives:** Identify neuromuscular risk factors for non-contact knee injury, using a systematic review and meta-analysis, to inform the development of preventive strategies.

**Methods:** Medline, Web of Science and SCOPUS were searched from inception until November 2020. Prospective and nested case-control studies that analysed baseline neuromuscular characteristics as potential risk factors for subsequent non-contact knee injuries were included. Two reviewers independently appraised methodological quality using a modified Newcastle–Ottawa Scale. Meta-analysis was performed where appropriate, with standardised mean differences calculated for continuous scaled data.

**Results:** Seventeen studies were included, comprising baseline data from 5,584 participants and 415 non-contact knee injuries (heterogeneous incidence = 7.4%). Protocols and outcome measures differed across studies, limiting data pooling. Twenty-one neuromuscular variables were included in the meta-analysis. Three were identified as risk factors. For patellofemoral pain, among military recruits: reduced non-normalised quadriceps strength at 60º/s (SMD −0.66; 95% CI −0.99, −0.32); reduced quadriceps strength at 240º/s (normalised by body mass) (SMD −0.53; CI −0.87, −0.20). For PFP/ACL injury among female military recruits: reduced quadriceps strength at 60º/s (normalised by body mass) (SMD −0.50; CI −0.92, −0.08).

**Conclusions:** Quadriceps weakness is a risk factor for PFP among military recruits, and for PFP/ACL injury among female military recruits. However, the effect sizes are small, and the generalisability of these findings is limited. The effectiveness of quadriceps strengthening interventions for preventing PFP and ACL injury merits evaluation in prospective randomised trials.

## INTRODUCTION

High rates of knee injury have been reported among physically active populations, including team sports athletes, runners, and military personnel.^1, 2^ Acute knee injuries often require surgical intervention,^3^ and chronic knee injuries frequently persist, despite evidence-based management.^4, 5^ Consequently, knee injuries result in more time loss than injuries to other lower limb regions.^6^ In the longer term, both acute and chronic knee injuries increase the risk of knee osteoarthritis and other knee-related symptoms, reducing quality of life.^7, 8, 9^ Prevention of knee injuries is critical to the physical and psychological health of physically active people, and success in competitive sport.

The ‘sequence of prevention’ model (Figure 1) provides a guide to developing injury prevention strategies.^10^ Knee injuries have both a high incidence and severity (Stage 1).^1-9^ Among potential risk factors, neuromuscular variables are particularly relevant to prevention, as they can be quantified using valid and reliable tests and modified through clinical intervention. For example, strength can be assessed by handheld dynamometry,^11^ and can be enhanced by resistance training.^12^ In contrast, other potential risk factors may require specialist expertise or technology to assess (e.g. bone anatomy), or may be unmodifiable (e.g. age, sex, previous injury).^12^ Identifying neuromuscular risk factors for knee injury facilitates understanding of the mechanisms of injury (Stage 2), and provides targets for modification by preventive interventions (Stage 3), the effectiveness of which can then be evaluated in clinical trials (Stage 4).

**Figure 1:**
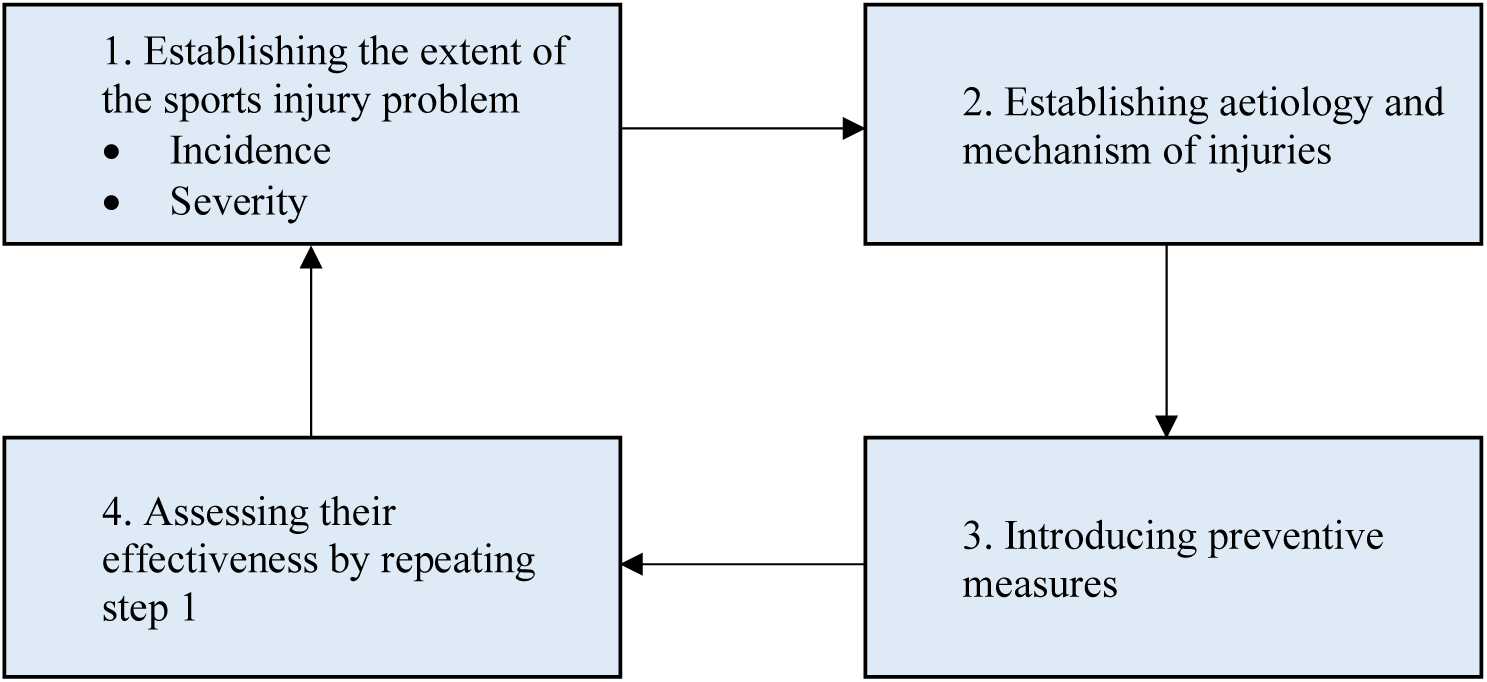
The sequence of prevention of sports injuries^10^.

Prospective study designs are best placed to identify possible risk factors for subsequent injury, when compared with case-control or cross-sectional study designs that are unable to eliminate or control for factors that associated with existing injury or pain. Previous systematic reviews of risk factors for knee injuries have identified quadriceps weakness as a risk factor for PFP among military recruits, and increased hip abduction strength as a risk factor for PFP among adolescents.^18^ However, rather than studying knee injuries in general, these reviews focused on specific diagnoses,^18, 19^ populations,^20^ or activities,^21^ reducing the number of eligible studies, and the number of variables for which data could be pooled for meta-analysis. Data syntheses have also been limited by the small number of high-quality prospective studies. For instance, Chester *et al*^*16*^ identified only one study of VMO activation timing as a risk factor for PFP that was of sufficient quality for inclusion. Thus, the extent to which neuromuscular variables can predict future knee injury remains unclear.

The aim of the present review was to synthesise evidence on neuromuscular risk factors for non-contact knee injuries, to inform the development of preventive interventions. To maximise the number of neuromuscular variables available for meta-analysis, we included prospective studies of risk factors for any type of non-contact knee injury, in any population, with exposure to any type of activity.

## METHODS

### Protocol and registration

The PRISMA checklist^22^ was consulted prior to the review and all items were completed.

### Eligibility criteria

**Table 1:**
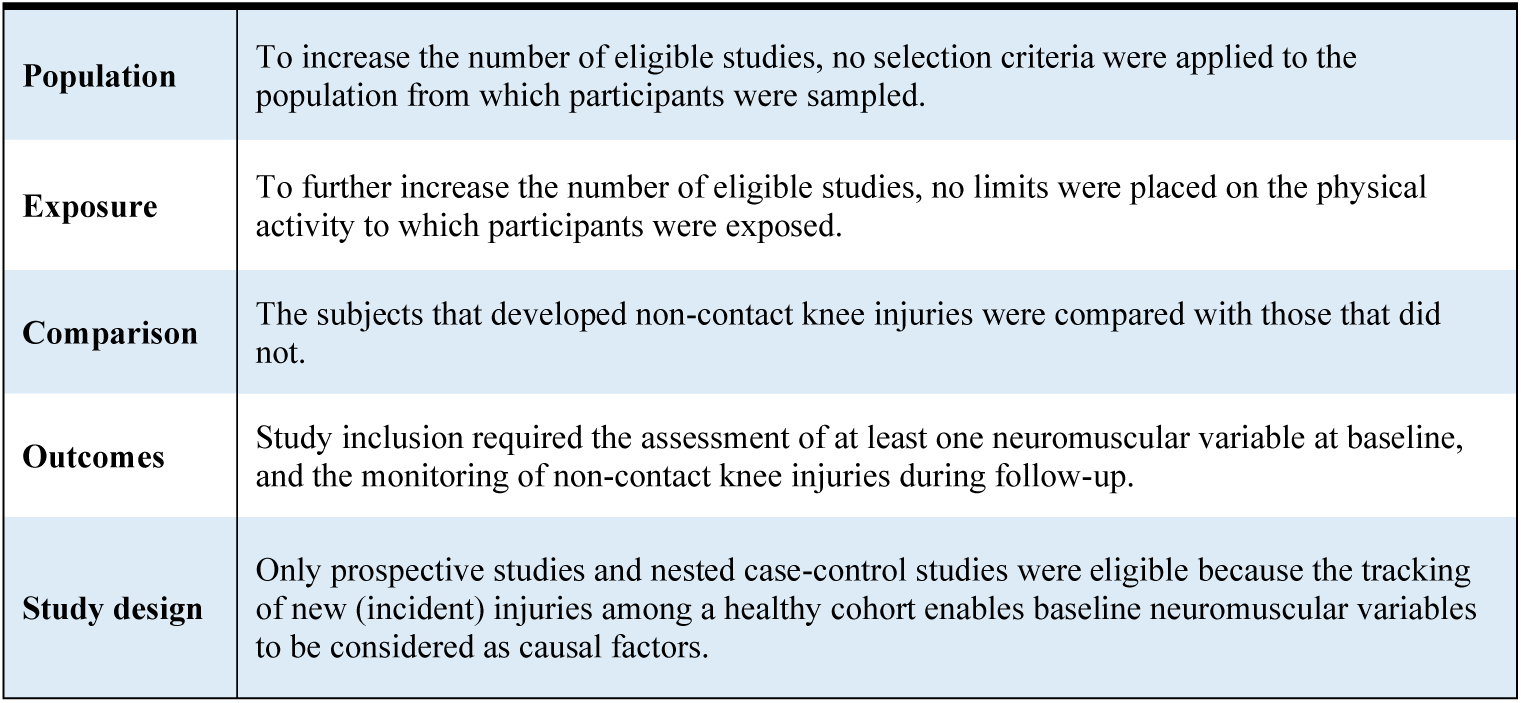
Eligibility criteria.

|  |  |
| --- | --- |
| **Population** | To increase the number of eligible studies, no selection criteria were applied to the population from which participants were sampled. |
| **Exposure** | To further increase the number of eligible studies, no limits were placed on the physical activity to which participants were exposed. |
| **Comparison** | The subjects that developed non-contact knee injuries were compared with those that did not. |
| **Outcomes** | Study inclusion required the assessment of at least one neuromuscular variable at baseline, and the monitoring of non-contact knee injuries during follow-up. |
| **Study design** | Only prospective studies and nested case-control studies were eligible because the tracking of new (incident) injuries among a healthy cohort enables baseline neuromuscular variables to be considered as causal factors. |

### Information sources

Medline, Scopus and Web of Science online databases were searched on 25^th^ November 2020, using the search strategy described in Table 2.

**Table 2:**
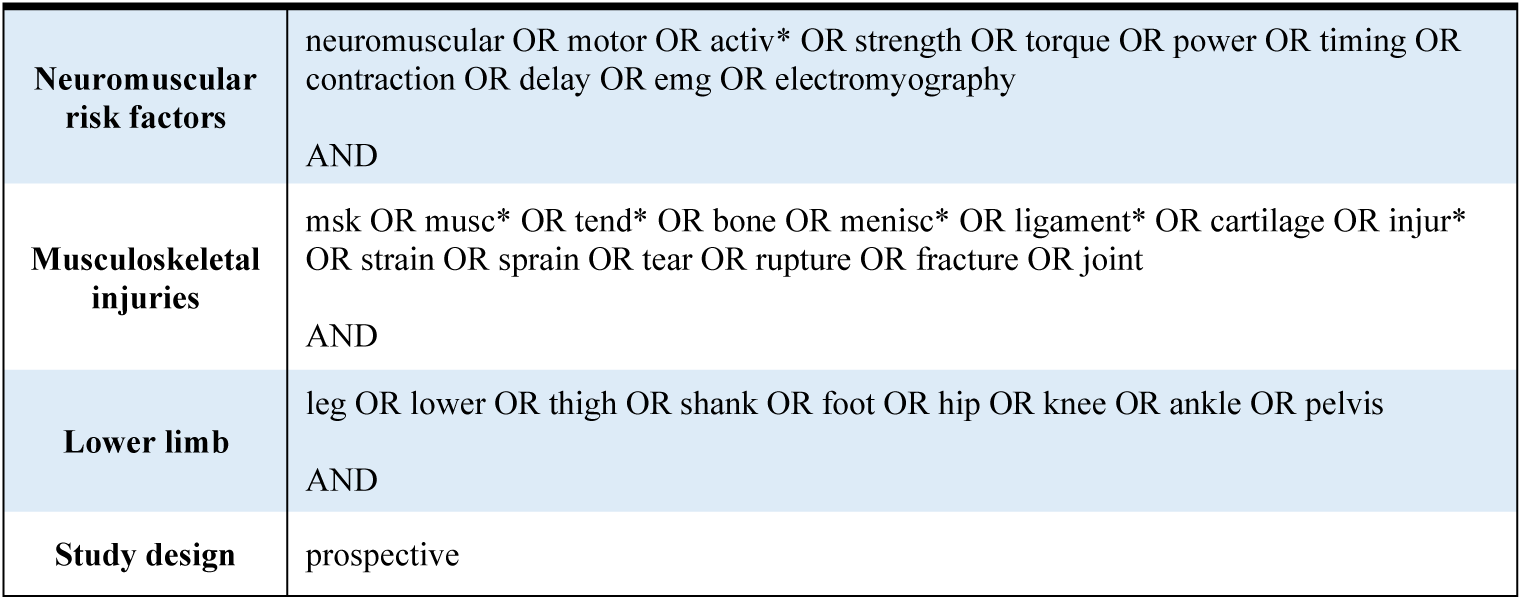
Search Strategy.

### Search

### Study selection

Children and older individuals are susceptible to growth- and frailty-related injuries, respectively. Thus, studies were excluded if participants had a mean age of <16 or >50 years. Studies were excluded if they did not distinguish between contact and non-contact injuries. However, studies of overuse injuries were not required to explicitly state that these had occurred via non-contact mechanisms.

Two authors (BM and BK) exported all articles identified by the search strategy to Mendeley Desktop (Version 1.19.4, Elsevier, New York, New York, USA). Articles were uploaded into Rayyan^23^ (Version 0.1.0, QCRI, Doha, Qatar) and blind screened (BM and BK) for relevance to non-contact knee injury. Once both authors had applied the selection criteria to all articles, blinding was removed. The full texts of the remaining articles were then screened to confirm their eligibility. In addition, the reference lists of articles included in the review, and of previous systematic reviews of risk factors for knee injuries, were hand-searched. Any conflicting selection decisions were discussed and resolved by the two authors. Reasons for excluding articles after full-text screening were recorded.

### Data collection process

Data related to study characteristics were extracted and entered on spreadsheets using Microsoft Excel (Version 16.35, Microsoft, Redmond, Washington, USA). When studies did not report all potentially relevant data, the corresponding author was emailed, and missing data requested.

Means and standard deviations for measures of strength, flexibility, balance and muscle activation were extracted. When numerical values were not available, these were obtained using graph digitiser software (WebPlotDigitizer, Version: 4.2, San Francisco, California, USA).

### Data items

Data were extracted on participants (population, number, baseline injury history); exposure (duration of follow-up period, type of physical activity, training requirements); baseline neuromuscular variables (summary measures, assessment protocols); knee injuries recorded during follow-up; and key findings.

### Risk of bias in individual studies

Because the review comprised prospective and nested case-control studies, methodological quality was independently assessed by two reviewers (BM and BK) using a modified Newcastle-Ottawa scale (NOS).^24^ NOS Criterion 5 relates to the comparability of a study’s cohorts. In the present review, this was modified to apply to the sexes of the participants, and the male-to-female ratio in the injured and uninjured groups, to account for sex differences in neuromuscular measures at baseline. A study was awarded one point for each criterion that it satisfied. In total, the modified NOS contained 8 categories relating to methodological quality. Each study was given a score, with a maximum of 8 points. Low- , moderate- and high-quality studies were defined as having scores of ≤4, 5-6, and 7-8 points respectively. Initial decisions were recorded and used to calculate inter-rater reliability. Conflicting decisions were discussed, and the authors made consensus decisions.

### Summary measures

The principal summary measures were the means and standard deviations (SDs) for baseline neuromuscular variables in the injured and uninjured groups. These were used to calculate standardised mean differences (SMDs) and 95% confidence intervals. Alternative summary measures included medians, risk ratios and odds ratios.

### Synthesis of results

Statistical analysis was performed using Review Manager (RevMan, [Computer program]. Version 5.3.5. Copenhagen: The Nordic Cochrane Centre, The Cochrane Collaboration, 2014).^25^ Means and SDs were extracted for continuous scaled variables and used to calculate standardised mean differences (SMDs) with 95% confidence intervals (CIs). Alternative summary measures (e.g. medians) were extracted and described in the text but not included in the meta-analysis.

Following the guidelines outlined by van Tulder *et al*,^26^ the level of evidence depended on the number and quality of studies that investigated each variable, and the homogeneity (consistency) of the direction and magnitude of its effect across different studies (Table 4).

**Table 3:** Selection Criteria.

|  |  |
| --- | --- |
| **Inclusion criteria** | 1. Prospective or nested case-control study design.<br>2. Participants were healthy (uninjured) at baseline.<br>3. Participants were exposed to physical activity of any type during the follow-up period.<br>4. At least one neuromuscular characteristic (potential risk factor) was assessed at baseline.<br>5. Recorded incident non-contact injuries to the lower limb (knee) during the follow-up period. |
| **Exclusion criteria** | 1. Mean age of participants was <16 years or >50 years at baseline, or, if no mean age was provided, participants were described as ‘child’, ‘youth’ or ‘older adults’.<br>2. Did not distinguish between contact injuries and non-contact injuries during data collection or statistical analysis.<br>(3. The specific location of incident injuries did not include the knee or a structure of the knee) |

**Table 4:** Levels of Evidence.

|  |  |
| --- | --- |
| **Strong evidence** | Statistically homogeneous findings across three or more studies, including a minimum of two high-quality studies. |
| **Moderate evidence** | Statistically homogeneous findings across two or more low- or moderate-quality studies.<br>OR<br>Statistically heterogeneous findings across two or more studies, at least one of which is high-quality. |
| **Limited evidence** | Statistically heterogeneous findings from one high-quality study.<br>OR<br>Statistically heterogeneous findings from two or more moderate- or low-quality studies. |
| **Very limited evidence** | Findings from one moderate- or low-quality study. |

Neuromuscular variables were categorised into domains of strength, flexibility, balance and muscle activation. Within each domain are specific characteristics, such as ‘quadriceps strength’, or ‘quadriceps flexibility’. If studies assessed a variable using different protocols or outcome measures, data were only pooled for meta-analysis if the respective methods were deemed sufficiently similar. For example, ‘concentric strength’ was not deemed similar enough to ‘isometric strength’ for data from these measures to be pooled.

A random effects model was used to account for between-study variation in participant characteristics and methods, and for the small number of studies. This reduced the probability of a type 1 error.^27^ Calculated individual and pooled SMDs were categorised as small (0.5-0.79), medium (0.80–1.19) or large (≥1.2). We used higher SMD thresholds than those proposed by Cohen,^28^ to reduce the probability that variables would be falsely labelled as risk factors (type 1 errors).^29^ Thus, in the present review, to be labelled as a risk factor, a variable had to have been investigated in two or more studies, have a pooled SMD ≤-0.5, and an upper confidence interval limit ≤0.

### Additional analyses

Because knee injury rates can differ between males and females,^30, 31, 32^ wherever possible, sex-specific data were extracted and analysed in male and female subgroups.

Similarly, wherever possible, data were pooled by specific population (e.g. military recruits) and analysed as subgroups.

## RESULTS

### Study selection

The initial ‘lower limb’ search identified 12,694 titles and abstracts. Of these, 12,588 articles were excluded as not relevant to the research question. After the aim of the review was updated (see Methods), articles unrelated to knee injury were excluded. Of the 54 articles that remained, 17 met inclusion criteria for the review, of which 10 were included in the meta-analysis (Figure 2).

**Figure 2:**
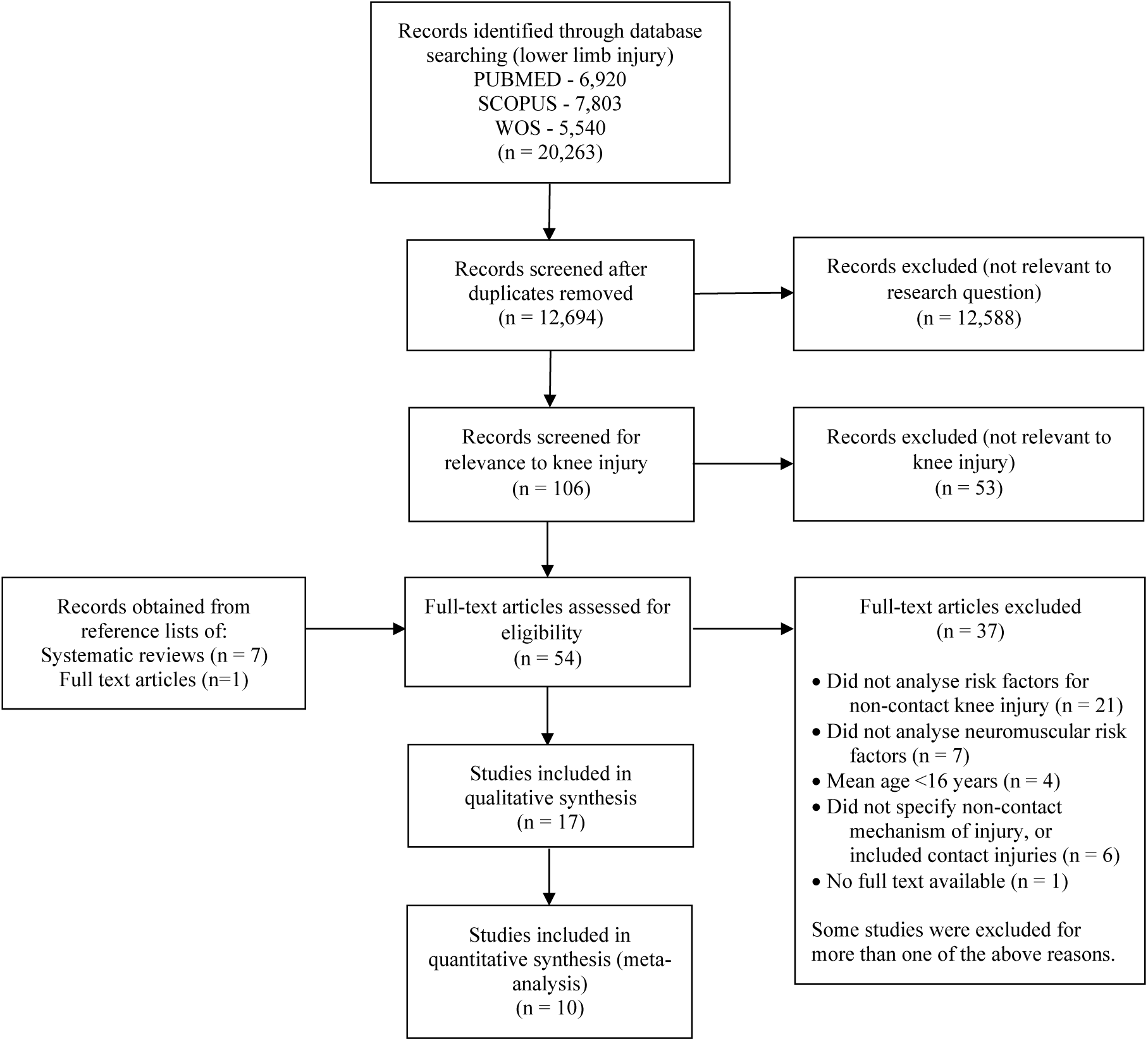
PRISMA search flowchart.

### Study characteristics

Thirteen studies^33, 34, 35, 36, 37, 38, 39, 40, 43, 44, 45, 47, 48^ assessed strength, four^41, 46, 47, 49^ assessed muscle activation, three^44, 47, 48^ assessed flexibility and two^42, 47^ assessed balance.

Nine studies^33, 35, 37, 38, 40, 43, 45, 46, 47^ investigated risk factors for patellofemoral pain (PFP). Six^35, 38, 40, 41, 43, 49^ investigated non-contact ACL injuries. One^48^ investigated patellar tendinopathy, and one^34^ investigated overuse knee injuries.

Five distinct populations were studied: military recruits,^33, 35, 38, 44, 45, 46^ adolescents,^34, 37, 39, 47, 48^ elite female athletes,^41, 42, 49^ recreational runners,^40, 43^ and club-level athletes.^26^

Seven studies^33, 36, 37, 40, 44, 47, 48^ recruited participants of both sexes, seven^34, 35, 39, 41, 42, 43, 49^ recruited only females, and three^38, 45, 46^ recruited only males.

Follow-up periods ranged in duration from 6 weeks to 8 years (median 12 months).

### Risk of bias within studies

Six studies^35, 43, 45, 46, 47, 48^ were deemed high quality (HQ), ten^33, 34, 36, 37, 38, 39, 40, 41, 42, 44^ were deemed moderate quality (MQ), and one^49^ was deemed low quality (LQ) (Table 6). The mean percentage agreement between the two authors for quality ratings was 90.25% (range 76-100%), indicating a high inter-rater reliability. The lowest rates of agreement (76%) were for Criteria 3 and 5. No study satisfied Criterion 2 (inclusion of a non-exposed control group).

**Table 5:**
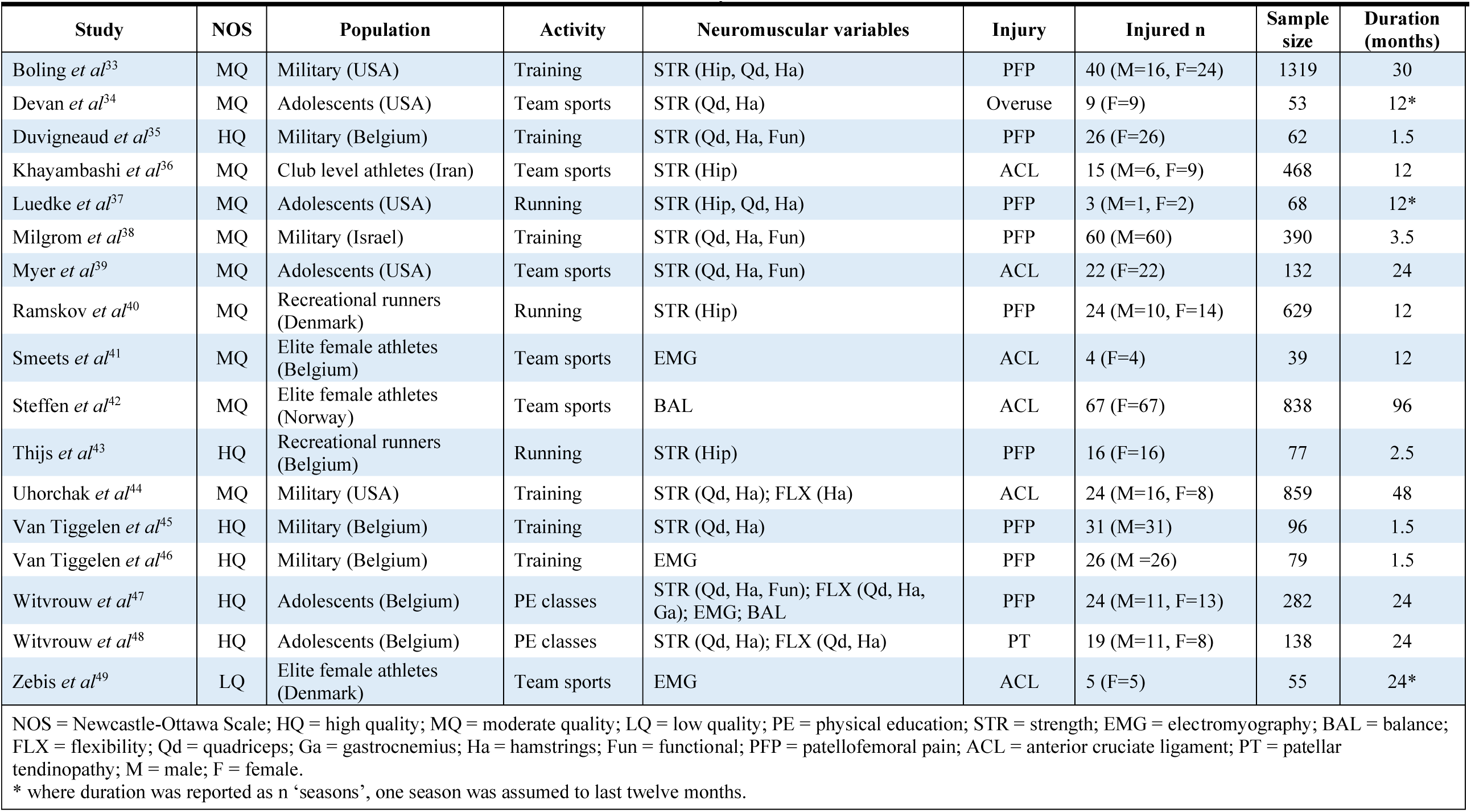
Study Characteristics.

**Table 6:** Modified Newcastle-Ottawa Scale consensus decisions for individual studies and percentage agreement.

| Criterion | Selection |  |  |  | Comparability | Outcomes |  |  | Score (/8) |
| --- | --- | --- | --- | --- | --- | --- | --- | --- | --- |
|  | The cohort was truly representative of the physically active population being investigated. | A non-exposed control group was included to determine the natural incidence of injury. | The type and amount of physical activity to which subjects were exposed was clearly described. | The injury being investigated was not present in any subject at the start of the study. | The study only included participants of one sex, or adequately controlled for sex during statistical analysis. | A clear definition of injury was provided, and the injury documentation process was clearly described. | Follow-up time was clearly stated and matched the duration of the exposure. | ≥80% of subjects were successfully followed up. |  |
| Boling *et al*<sup>33</sup> | Y | N | Y | Y | N | Y | Y | Y | 6 |
| Devan *et al*<sup>34</sup> | Y | N | N | Y | Y | Y | N | Y | 5 |
| Duvigneaud *et al*<sup>35</sup> | Y | N | Y | Y | Y | Y | Y | Y | 7 |
| Khayambashi *et al*<sup>36</sup> | Y | N | N | Y | N | Y | Y | Y | 5 |
| Luedke *et al*<sup>37</sup> | Y | N | Y | Y | N | Y | Y | Y | 6 |
| Milgrom *et al*<sup>38</sup> | Y | N | N | Y | Y | Y | Y | Y | 6 |
| Myer *et al*<sup>39</sup> | Y | N | N | Y | Y | Y | Y | Y | 6 |
| Ramskov *et al*<sup>40</sup> | Y | N | Y | Y | Y | Y | Y | N | 6 |
| Smeets *et al*<sup>41</sup> | Y | N | N | Y | Y | Y | N | Y | 5 |
| Steffen *et al*<sup>42</sup> | Y | N | N | Y | Y | Y | Y | Y | 6 |
| Thijs *et al*<sup>43</sup> | Y | N | Y | Y | Y | Y | Y | Y | 7 |
| Uhorchak *et al*<sup>44</sup> | Y | N | Y | Y | Y | Y | Y | N | 6 |
| Van Tiggelen *et al*<sup>45</sup> | Y | N | Y | Y | Y | Y | Y | Y | 7 |
| Van Tiggelen *et al*<sup>46</sup> | Y | N | Y | Y | Y | Y | Y | Y | 7 |
| Witvrouw *et al*<sup>47</sup> | Y | N | Y | Y | Y | Y | Y | Y | 7 |
| Witvrouw *et al*<sup>48</sup> | Y | N | Y | Y | Y | Y | Y | Y | 7 |
| Zebis *et al*<sup>49</sup> | Y | N | N | Y | Y | N | N | Y | 4 |
| Initial percentage agreement (%) | 100 | 100 | 76 | 100 | 76 | 100 | 82 | 88 | - |

Y = Yes; N = No; A highlighted box indicates that discussion between the two assessors was required to resolve a conflicting decision. Only consensus decisions are shown.

### Strength

All studies that investigated concentric and eccentric strength used isokinetic dynamometry (IKD). Studies that investigated isometric strength used handheld dynamometry (HHD). Studies either reported absolute (non-normalised) strength, relative strength (normalised by body mass), or both. Absolute strength data were pooled separately from relative strength data in the meta-analysis.

#### Knee extension (quadriceps) strength

Moderate evidence (two HQ studies^35, 45^) of small effect indicates absolute concentric knee extension weakness at 60º/s is a risk factor (SMD −0.66; CI −0.99, −0.32; I^2^ = 0%) for patellofemoral pain (PFP) in military recruits, but not at 240º/s (SMD −0.49; CI −0.85, −0.12; I^2^ = 17%) (Figures 3.1.1 and 3.1.2). Moderate evidence (three HQ studies, ^35, 45, 48^ one MQ study^44^) indicates that concentric knee extension weakness at 60º/s, normalised to body mass, is not a risk factor (SMD −0.33; CI −0.64, −0.02; I^2^ = 45%) for non-contact knee injury at 60º/s (Figure 3.1.3). This finding remained true (SMD −0.44; CI −0.75, −0.13; I^2^ = 30%) when data were pooled for military recruits only^35, 44, 45^ (Figure 3.1.4).

Moderate evidence (three HQ studies^35, 45, 48^) indicates that concentric knee extension weakness at 240º/s, normalised to body mass, is not a risk factor (SMD −0.35; CI −0.74, 0.03; I^2^ = 48%) for non-contact knee injury (Figure 3.1.5), but moderate evidence (two HQ studies^35, 45^) of small effect indicates that it is a risk factor (SMD −0.53; CI −0.87, −0.20; I^2^ = 0%) for PFP when data were pooled for military recruits only (Figure 3.1.6).

Sex-specific data could only be pooled for concentric knee extension strength at 60º/s, normalised by body mass. Moderate evidence (one HQ study,^45^ one MQ study^44^) indicates that it is not a risk factor (SMD −0.19; CI −0.98, 0.59; I^2^ = 82%) for non-contact knee injury in male military recruits (Figure 3.1.7). Moderate evidence (one HQ study,^35^ one MQ study^44^) of small effect indicates that it is a risk factor (SMD −0.50; CI −0.92, −0.08; I^2^ = 0%) for non-contact knee injury in female military recruits (Figure 3.1.8).

Limited evidence (two MQ studies^33, 38^) indicates that isometric knee extension weakness, normalised to body mass, is not a risk factor (SMD −0.25; CI −0.74, 0.25; I^2^ = 82%) for PFP in military recruits (Figure 3.1.9). Very limited evidence (one MQ study^38^) indicates that absolute isometric knee extension weakness is not a risk factor (SMD 0.28; CI 0.01, 0.56) for PFP in military recruits. Luedke *et al*^37^ (MQ) found that adolescent runners in the weakest third for knee extension strength had a higher incidence of PFP than those in the strongest third (chi-square = 6.562; P = 0.046).

Limited evidence (one HQ study^35^) indicates that absolute eccentric quadriceps strength at 30º/s is not a risk factor (SMD −0.15; CI −0.66, 0.35) for PFP in military recruits. Very limited evidence (one MQ study^34^) indicates that eccentric quadriceps strength at 60º/s, normalised by body mass, is not a risk factor (SMD −0.13; CI −0.55, 0.28) for ACL injury in military recruits.

#### Knee flexion (hamstrings) strength

Moderate evidence (two HQ studies^35, 45^) indicates that absolute concentric knee flexion weakness is not a risk factor for PFP in military recruits at 60º/s (SMD −0.09; CI −0.42, 0.24; I^2^ = 0%) (Figure 3.2.1) or at 240º/s (SMD −0.10; CI −0.43, 0.22; I^2^ = 0%) (Figure 3.2.2).

Moderate evidence (one HQ study,^48^ one MQ study^44^) indicates that concentric knee flexion weakness at 60º/s, normalised by body mass, is not a risk factor (SMD −0.16; CI −0.48, 0.15; I^2^ = 0%) for non-contact knee injury (Figure 3.2.3).

Eccentric knee flexion strength data could not be pooled for meta-analysis. Limited evidence (one HQ study^35^) indicates that absolute eccentric hamstrings strength at 30º/s is not a risk factor for PFP in military recruits (SMD −0.49; CI -1.00, 0.02). Very limited evidence (one MQ study^44^) indicates that eccentric hamstrings strength at 60º/s, normalised by body mass, is not a risk factor (SMD 0.01; CI −0.40, 0.43) for ACL injury in military recruits.

Isometric knee flexion strength data could not be pooled for meta-analysis because only one study reported means and standard deviations. Very limited evidence (one MQ study^33^) indicates that isometric knee flexion strength is not a risk factor (SMD −0.40; CI −0.72, −0.08) for PFP in military recruits. Luedke *et al*^37^ (MQ) found that adolescent runners in the weakest third for knee flexor strength had a higher incidence of PFP than those in the strongest third (chi-square = 6.140; P = 0.046).

#### Hamstrings-to-quadriceps ratio

Limited evidence (one HQ study^35^) of small effect indicates that a high hamstrings-to-quadriceps ratio is a risk factor for PFP in military recruits at both 60º/s (SMD 0.6; CI 0.08, 1.12) and 240º/s (SMD 0.59; CI 0.08, 1.11). One MQ study^34^ found that prevalence of overuse knee injuries was greater in athletes with H:Q ratios below the normal range at both 60º/s and 300º/s (p = 0.004). One MQ study^39^ found that the median hamstrings to quadriceps ratio at 300º/s was lower in adolescents that developed ACL injuries compared with those that remained uninjured.

#### Hip strength

Moderate evidence (one HQ study,^43^ two MQ studies^33, 36^) indicates that isometric hip abduction weakness, normalised by body mass (SMD −0.46; CI −0.86, −0.06; I^2^ = 57%), and isometric hip external rotation weakness, normalised by body mass (SMD −0.38; CI −0.79, 0.04; I^2^ = 61%) are not risk factors for non-contact knee injury (Figures 3.3.1 and 3.3.2).

Moderate evidence (one HQ study,^43^ one MQ study^33^) indicates that isometric hip internal rotation weakness, normalised by body mass (SMD −0.11; CI −0.49, 0.26; I^2^ = 36%), and isometric hip extension weakness (SMD −0.23; CI −0.50, 0.05; I^2^ = 0%), normalised by body mass, are not risk factors for non-contact knee injury (Figures 3.3.3 and 3.3.4).

Hip flexion and hip adduction strength were each assessed only in one study. Limited evidence (one HQ study^43^) indicates that isometric hip flexion (SMD 0.28; CI −0.27, 0.83) and isometric hip adduction (SMD −0.32; CI −0.87, 0.23) strength are not risk factors for PFP in runners.

#### Functional strength

Moderate evidence (one HQ study,^47^ one MQ study^38^) indicates that trunk flexion weakness, assessed by sit-ups, is not a risk factor (SMD −0.06; CI −0.29, 0.17; I^2^ = 0%) for PFP (Figure 3.4).

**Figure 3.1:**
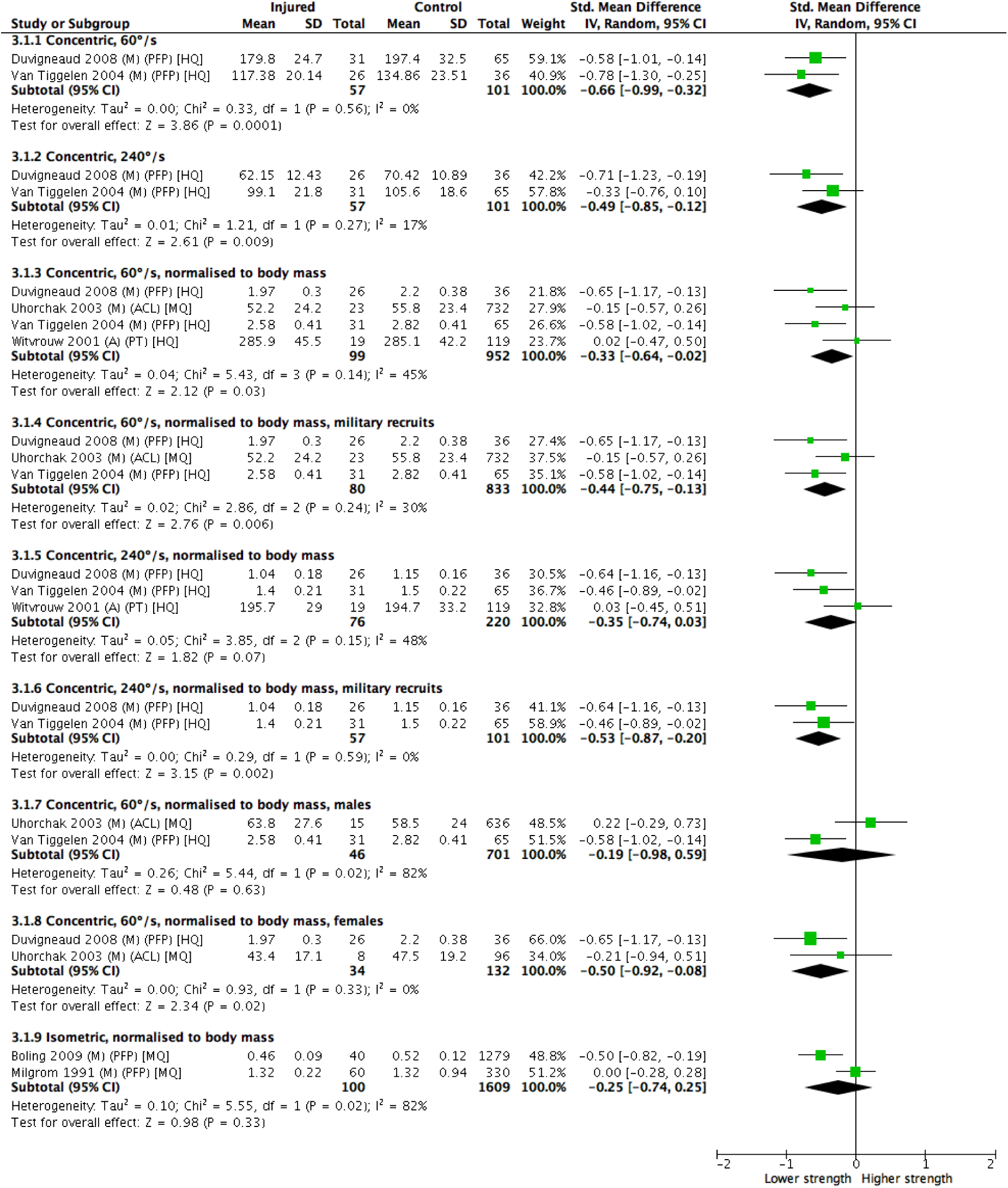
Quadriceps strength. M = military; A = adolescents; PFP = patellofemoral pain; ACL = anterior cruciate ligament; PT = patellar tendinopathy; HQ = high quality; MQ = moderate quality.

**Figure 3.2:**
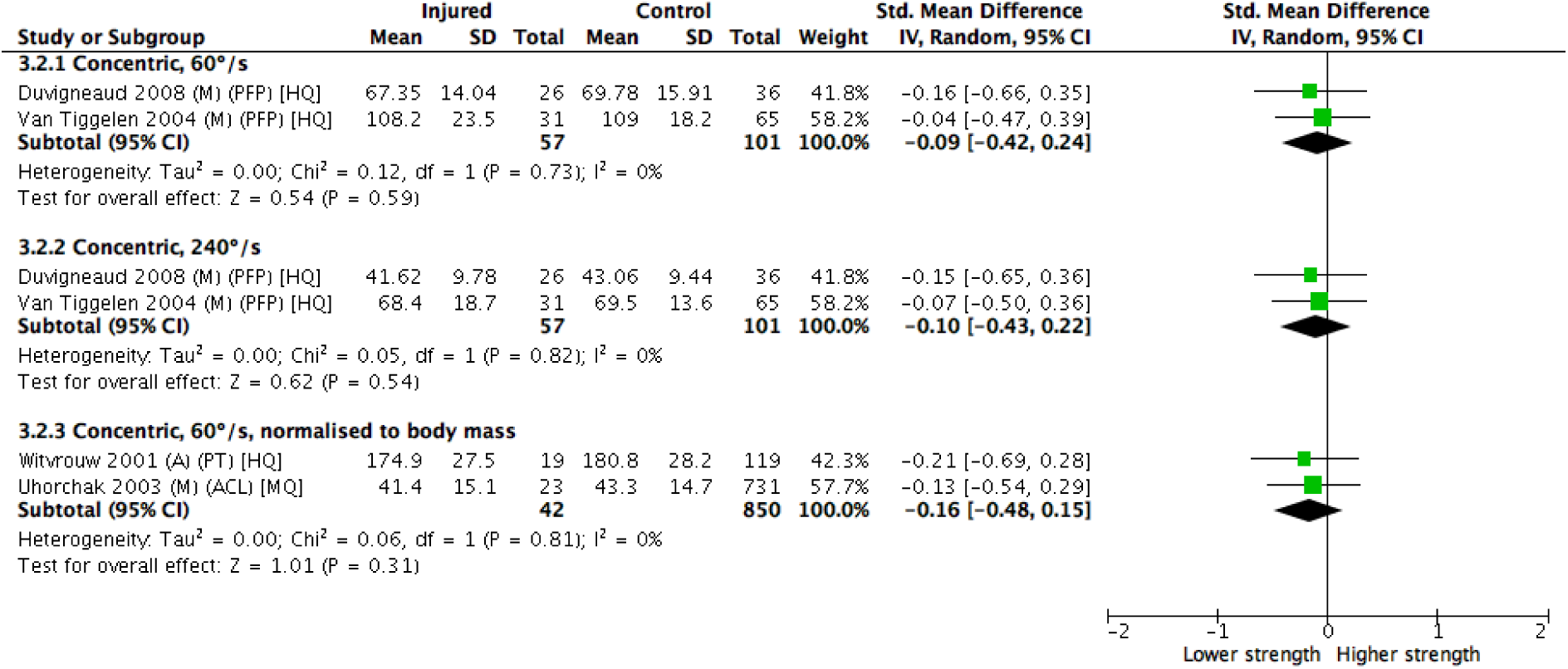
Hamstrings strength. M = military; A = adolescents; PFP = patellofemoral pain; PT = patellar tendinopathy; ACL = anterior cruciate ligament; HQ = high quality; MQ = moderate quality.

**Figure 3.3:**
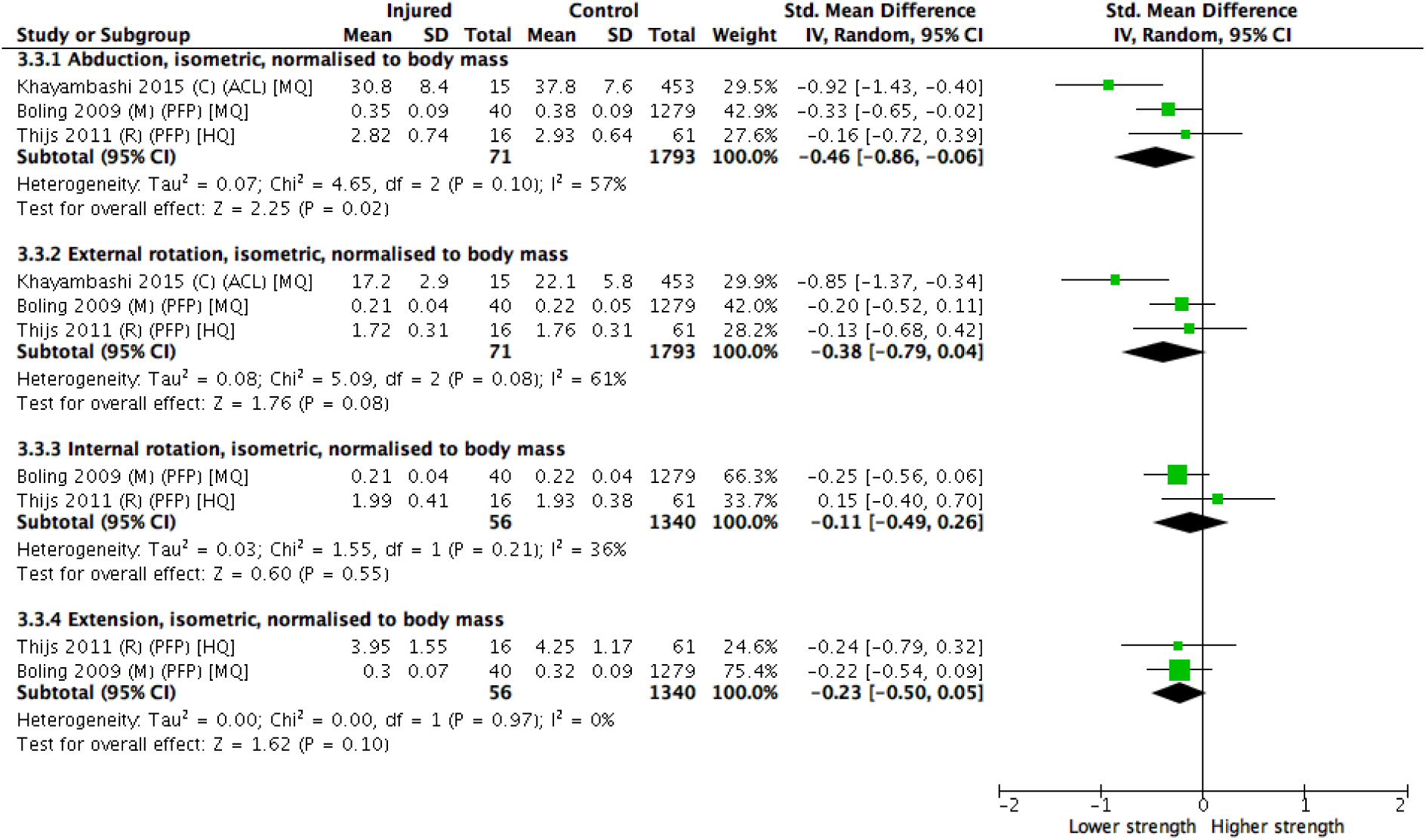
Hip strength. M = military; A = adolescents; PFP = patellofemoral pain; ACL = anterior cruciate ligament; PT = patellar tendinopathy; HQ = high quality; MQ = moderate quality.

**Figure 3.4:**
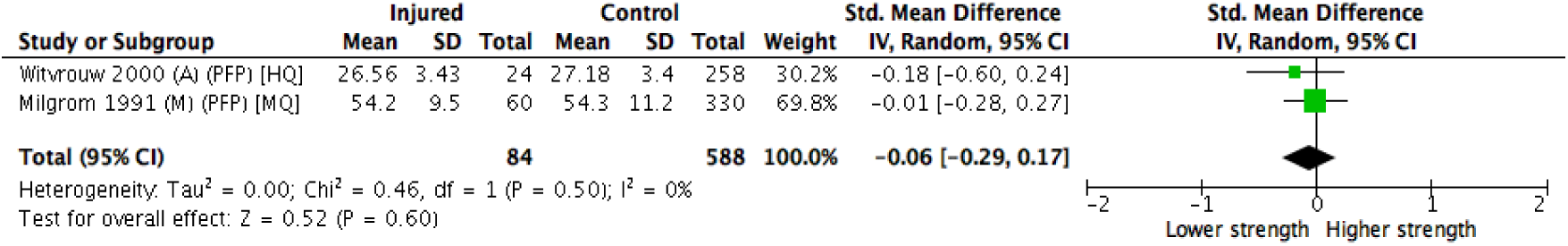
Trunk flexion strength. M = military; A = adolescents; PFP = patellofemoral pain; HQ = high quality; MQ = moderate quality.

Limited evidence (one HQ study^37^) of small effect indicates that lower vertical jump height is a risk factor (SMD −0.56; CI −0.98, −0.13) for PFP in adolescents. Myer *et al*^39^ (MQ) reported a median vertical jump height of 15.8cm in adolescents that sustained an ACL injury compared with 15.5cm in those that remained uninjured.

### Flexibility

Moderate evidence (two HQ studies^47, 48^) indicates that reduced quadriceps flexibility is not a risk factor (SMD -1.81; CI -4.46, 0.84; I^2^ = 98%) for non-contact knee injury in adolescents (Figure 4.1).

**Figure 4.1:**
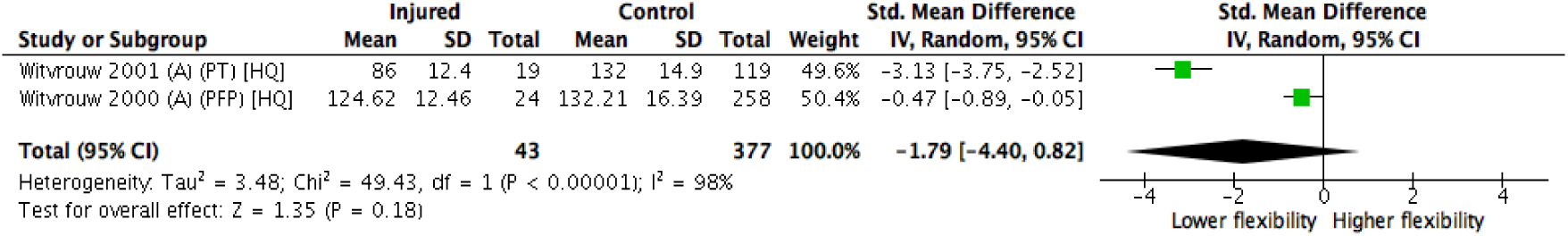
Quadriceps flexibility. A = adolescents; PT = patellar tendinopathy; PFP = patellofemoral pain; HQ = high quality.

**Figure 4.2:**
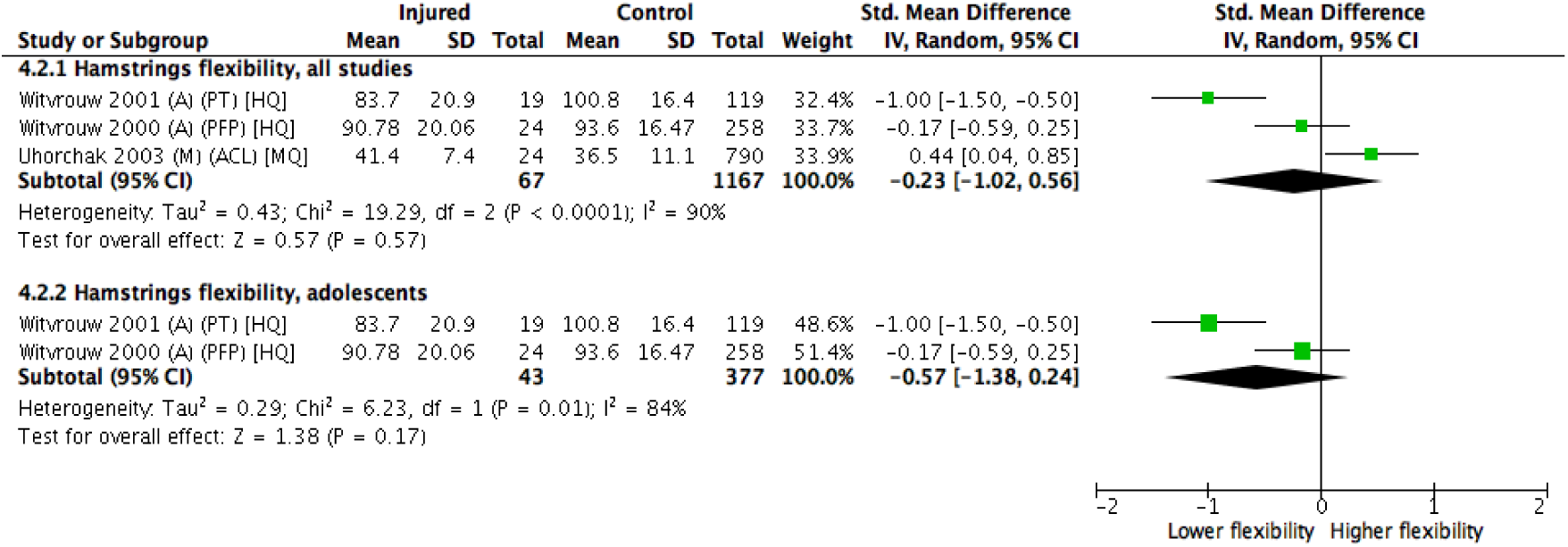
Hamstrings flexibility. A = adolescents; M = military recruits; PT = patellar tendinopathy; PFP = patellofemoral pain; ACL = anterior cruciate ligament; HQ = high quality; MQ = moderate quality.

Moderate evidence (two HQ studies,^47, 48^ one MQ study^44^) indicates that reduced hamstrings flexibility is not a risk factor (SMD −0.23; CI -1.02, 0.56; I^2^ = 90%) for non-contact knee injury (Figure 4.2.1). This remained true when data were pooled for adolescents only (small SMD −0.57; CI -1.38, 0.24; I^2^ = 84%) (Figure 4.2.2).

Limited evidence (one HQ study^47^) indicates gastrocnemius flexibility is not a risk factor (SMD - 0.48; CI −0.90, −0.06) for PFP in adolescents.

### Balance

The ‘balance platform test (static), 95% confidence ellipse’ in Steffen *et al*^42^ (MQ) was considered to be similar enough to the Flamingo balance test in Witvrouw *et al*^47^ (HQ) for data pooling. Moderate evidence (one HQ study,^47^ one MQ study^42^) indicates that reduced single leg balance is not a risk factor (SMD 0.17; CI −0.07, 0.41; I^2^ = 0%) for non-contact knee injury (Figure 5).

**Figure 5:**
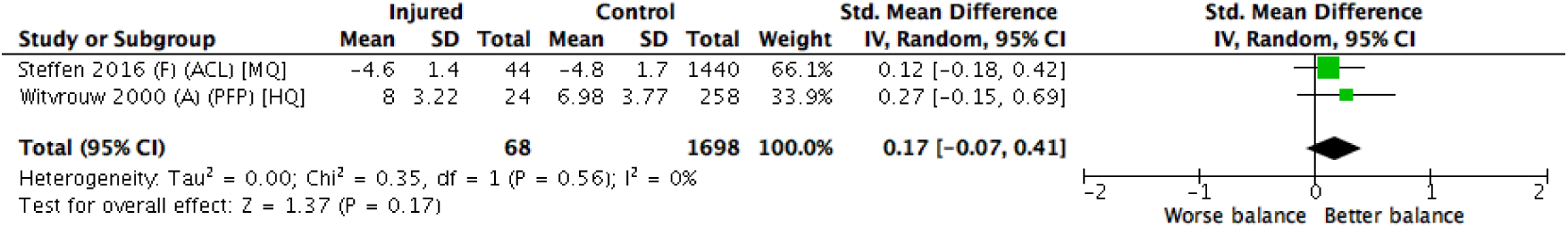
Single leg static balance. F = female athletes; A = adolescents; ACL = anterior cruciate ligament; PFP = patellofemoral pain; HQ = high quality; MQ = moderate quality.

### Muscle activation

Four studies assessed baseline muscle activation by electromyography. However, their methods were not similar enough for data to be pooled for meta-analysis.

Limited evidence (one HQ study^46^) of large effect indicates that delayed vastus medialis obliquus (VMO) activation in relation to vastus lateralis (VL) is a risk factor (SMD -1.58; CI -2.11, -1.05) for PFP in military recruits.

Limited evidence (one HQ study^47^) of small effect indicates that shortened VMO (SMD −0.50; CI - 0.92, −0.08) and VL (SMD −0.64; CI -1.06, −0.22) reflex response times are risk factors for PFP in military recruits.

Very limited evidence (one LQ study^49^) of large effect indicates that reduced semitendinosus activity (SMD -1.16; CI -2.1, −0.21) and increased vastus lateralis activity (SMD 2.30; CI 1.28, 3.31), prior to a cutting manoeuvre, are risk factors for ACL injury in elite female athletes.

Very limited evidence (one MQ study^41^) indicates that (hamstrings medialis, hamstrings lateralis), (vastus lateralis, hamstrings lateralis) and (hamstrings lateralis, vastus medialis) vector activations around peak loading and just before take-off are greater (P < 0.001) among female athletes that subsequently suffered ACL injuries compared with those that remained uninjured.

### Risk of bias across studies

Some authors were involved in multiple studies included in the present review. In particular, *Witvrouw, E*. was the first author of two articles,^47, 48^ and a co-author in four others.^35, 43, 45, 46^ Nevertheless, all of these studies were rated as either high (n=4) or moderate (n=2) quality.

## DISCUSSION

The aim of this systematic review was to identify neuromuscular risk factors for non-contact knee injuries. The key findings were that lower absolute concentric quadriceps strength at 60º/s, and lower quadriceps strength at 240º/s (normalised by body mass) were risk factors for patellofemoral pain (PFP) among military recruits; reduced concentric knee extension strength at 60º/s, normalised by body mass, was a risk factor for PFP and non-contact ACL injury, specifically among female military recruits. For each risk factor, the effect size (according to our thresholds) was small (-0.66, −0.53, and −0.50, respectively).

### Strength

Our findings are consistent with those from a previous systematic review,^18^ which identified the same two quadriceps weakness risk factors for PFP, after pooling data from the same two prospective studies of Belgian military recruits.^35, 45^ Although each study lasted only six weeks, there was a remarkably high incidence of PFP (41.9% and 32.3%, respectively). Studies of PFP among military recruits in other settings have reported much lower rates (3.0%^33^ and 15.4%^38^), despite longer follow-up periods (130 weeks and 15 weeks, respectively). This suggests that in the two studies in which we and Neal *et al*^18^ identified quadriceps weakness as a risk factor for PFP, the extrinsic training load was particularly high. In line with this, we found no evidence that quadriceps strength was a risk factor for PFP from studies in non-military cohorts.^48^ Thus, the generalisability of the findings is limited. Although only three strength variables in the meta-analysis met our risk factor criteria, subjects who developed injuries were weaker on every strength variable (Figures 3.1, 3.2, 3.3 and 3.4), with pooled SMDs ranging from −0.06 to −0.66.

No measure of hip strength was found to be a risk factor. This is surprising, given that proximal (hip) strengthening exercises are effective in the treatment of PFP^57^ and improve pain and function more than quadriceps rehabilitation.^57, 58^ However, the clinical benefits of proximal rehabilitation may result from other mechanisms than increased hip muscle strength.^57^ It is also possible that we were unable to detect an effect of hip strength because statistical power was limited, and subgrouping by population or sex was not possible. Although Neal *et al*^18^ identified *increased* hip abduction strength as a risk factor for PFP in adolescents, they suggested that this reflected an adaptation to increased physical activity, and that this accounted for the association.

### Flexibility

Quadriceps flexibility had the largest pooled effect size (large SMD -1.81) of any variable. This was driven by data from a single high quality study^48^ which reported a very large difference (SMD -3.18, CI -3.81, -2.58) in passive knee flexion range of motion (ROM) between adolescent physical education students that developed patellar tendinopathy (PT) (ROM 86.0º ± 12.4º) and controls (ROM 132.6º ± 14.9º). However, when these data were pooled with data from a study investigating PFP,^47^ quadriceps flexibility could no longer be considered a risk factor, as the pooled upper CI limit was positive (0.86). Nevertheless, reduced quadriceps flexibility may still be a risk factor for PT specifically, and given the large effect size, this relationship merits further investigation.

### Muscle activation

An alteration in the relative timing of the onset of vastus medialis obliquus (VMO) and vastus lateralis (VL) activation, which disrupts mediolateral patellar tracking, may underlie PFP.^59, 60, 61^ A previous systematic review^62^ found a trend towards delayed VMO activation in PFP subjects, but data synthesis was limited by between-study heterogeneity. Similarly, in this review, data from the two studies^46, 47^ that investigated VMO and VL activation could not be pooled for meta-analysis because of methodological differences. Nevertheless, both studies were of high quality and found that VMO activation was delayed in subjects that later developed PFP. Moreover, Van Tiggelen *et al*^46^ found that the larger the delay in VMO activation at baseline, the higher the risk of later PFP. It was reported that after completing military training, onset of VMO activation occurred after VL in all subjects that had developed PFP, but only a third of those who had not developed PFP. The latter finding raises the possibility that delayed VMO contraction may result from PFP, as well as being a causal factor.

### Balance

A previous systematic review^63^ concluded that balance training should be included in ACL injury prevention programmes. However, we identified only two studies^42, 48^ that assessed balance, and neither found associations between balance measures and subsequent knee injury (ACL and PFP, respectively).

### Clinical Implications

It has been argued that “screening tests to predict injury do not work” because they measure continuous variables which tend to be normally distributed (Figure 6).^50^ Hence, on the basis of a single test, even if a variable is significantly associated with increased injury risk, it is rarely possible to identify a cut-off point that is both sensitive and specific enough to justify risk stratification. Our findings support this view, as the variables that we found to be associated with non-contact knee injury had small effect sizes, and would not, in isolation, be strong predictors. However, prediction of knee injury may still be possible if different risk factors are integrated in multi-factorial models.^18, 51^ This approach has proven successful in other fields of medicine: for example, the Fracture Risk Assessment Tool (FRAX)^52, 53^ and the QRISK3-2018 calculator^54^ are used to predict the risk of osteoporotic fractures and cardiovascular events, respectively.

**Figure 6:**
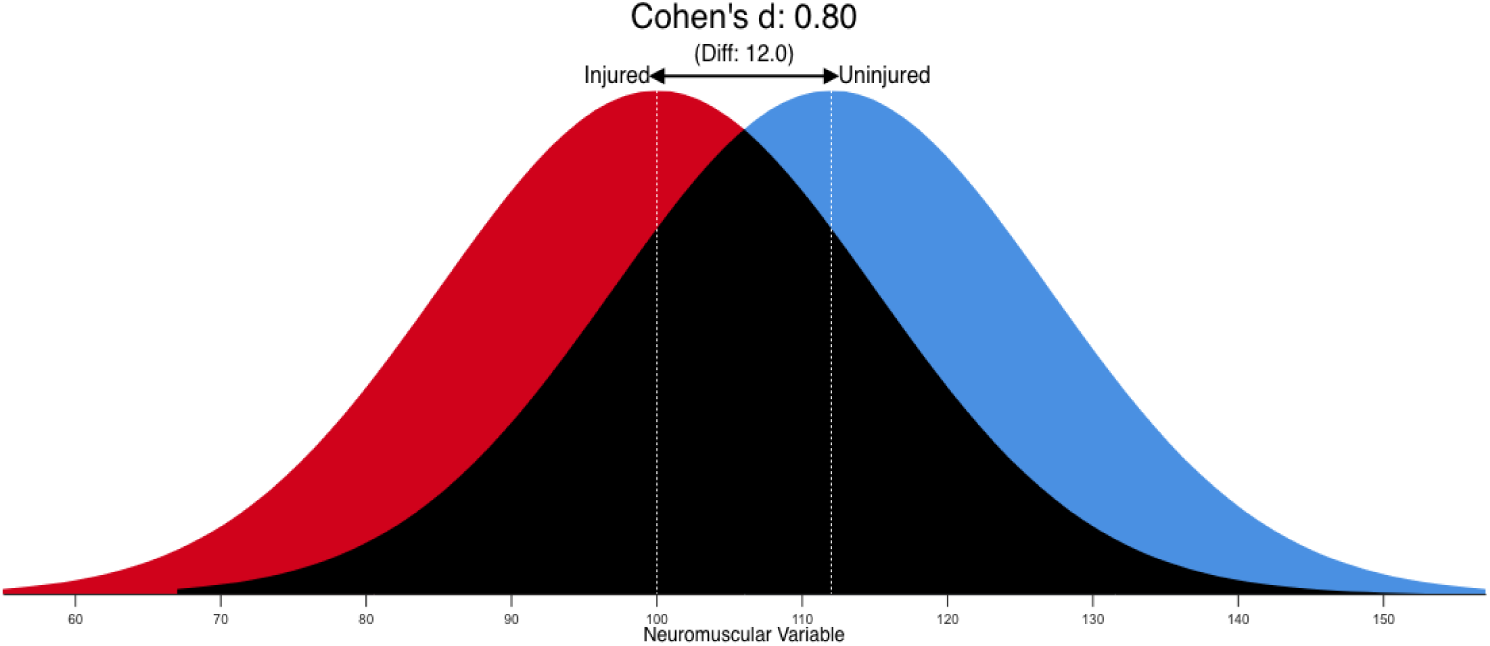
Illustration of the substantial (68.9%) overlap (black) between the values of injured (red) and uninjured (blue) subjects for a putative neuromuscular variable, even when the effect size (according to Cohen’s thresholds)^28^ is large (Cohen’s d = 0.8). Injured and control groups were assigned means of 100 and 112, respectively, with normal distributions, and SDs of 15. In this example, there is a 71.4% chance that a person picked at random from the injured group will have a lower score than a person picked at random from the control group.^29^

Testing protocols and outcome measures differed across studies, which reduced the number of variables with data that could be pooled for meta-analysis. This issue could be addressed by standardising protocols and measures across research groups within this field, as has proven successful in other research areas.^63, 64^

All studies in the present review involved subjects who were asymptomatic at baseline. However, the criteria used to exclude participants on the basis of their injury history differed greatly between studies, which may have affected the results. For instance, Uhorchak *et al*^44^ excluded volunteers with previous ACL injuries but Steffen *et al*^42^ did not, and found that a previous ACL injury tripled the odds for sustaining a new ACL injury. Previous musculoskeletal injuries are common in physically active individuals, so the exclusion of previously injured volunteers may result in an unrepresentative sample and an underpowered study. On the other hand, if subjects with previous injuries are to be included, it is unclear how long they should have been injury-free, and which types of previous injury are acceptable. The development of standardised participant selection criteria that take these considerations into account would facilitate future research in this area.

Some of the testing protocols used in the studies we examined have limited clinical applicability. For example, in team sports, the assessment of quadriceps strength using isokinetic dynamometry (IKD) may be too time-consuming to test multiple players, and teams may not have access to an IKD machine. Future research should employ more practical means of assessing neuromuscular characteristics. For example, handheld dynamometry represents a valid and reliable alternative to IKD.^11, 55, 56^

### Limitations

Not restricting our review to studies of a particular athletic population, physical exposure, or type of knee injury, allowed us to increase the number of neuromuscular variables available for meta-analysis, addressing a limitation of previous reviews. For example, pooling data from military recruits, club-level athletes and recreational runners permitted meta-analysis of hip abduction strength (Figure 3.3.1). However, this approach limits the clinical applicability of the findings to specific populations, and this could only be partially mitigated by population- and sex-specific subgrouping, due to the small number of studies (maximum four) from which data were pooled for each variable.

Applying conventional effect size thresholds would have resulted in more risk factors being identified. Many of these would have been very weak predictors of injury (Figure 6), but could represent areas for further research.

### CONCLUSIONS

#### Key findings

- Three measures of quadriceps strength were identified as risk factors for knee injury, but only from studies of military recruits.
- Large effect sizes were detected for reduced quadriceps flexibility and altered muscle activation, but only in single studies.
- No other neuromuscular variables were identified as risk factors for non-contact knee injury.

#### Clinical implications

- Standardisation of participant selection criteria, testing protocols and outcome measures across studies would enable future meta-analyses to examine more potential neuromuscular risk factors.
- A single measure of quadriceps strength is unlikely to be a strong predictor of knee injury, but might be useful when combined with other risk factors in a multifactorial model.
- The effectiveness of quadriceps strengthening interventions for prevention of PFP and ACL injury merits evaluation in randomised controlled trials.

## Data Availability

This was a systematic review and meta-analysis. Data from individual studies were obtained online.

